# ESOPHAGEAL ACID AND SYMPTOM FREQUENCY IN GERD PHENOTYPES: A QUADRANT-BASED ANALYSIS OF SUBJECT-LEVEL VARIATION

**DOI:** 10.1101/2025.06.29.25330512

**Authors:** Jerry D. Gardner, George Triadafilopoulos

**Affiliations:** Division of Gastroenterology and Hepatology, Stanford University School of Medicine, 420 Broadway Street, Pavilion D, 2nd Floor, Redwood City, CA 94063; Department of Gastroenterology, Hepatology and Nutrition, UT MD Anderson Cancer Center,100 Fellowship Drive, Conroe, TX 77384

## Abstract

**Background:** The Lyon Consensus identified distinct phenotypes of symptomatic GERD subjects: NERD, reflux hypersensitivity, and functional heartburn; however, the Consensus did not describe a relationship between esophageal acid exposure and symptom frequency.

**Objective:** The present analyses aim to examine this relationship in ways that capture the variation among individual GERD subjects.

**Design:** Records of symptoms and 24-hour esophageal pH from 60 subjects with a normal upper endoscopy were grouped using the Lyon criteria for the three phenotypes of GERD (20 subjects per phenotype). Interval esophageal acidity was calculated before each symptom from 24-hour pH recordings. The value for symptom frequency was plotted versus the corresponding value for esophageal acid for each subject on a graph divided into quadrants based on the median value for symptom frequency and esophageal acid from all 60 subjects. Thus, each subject was categorized as esophageal acid: symptom frequency as high: high; high: low; low: high or low: low.

**Results:** Subjects were distributed among all 4 quadrants, and each quadrant tended to cluster a specific Lyon Consensus phenotype (Chi-Square P=0.00018). There was a significant discordant relationship between esophageal acid and symptom frequency in 63% of subjects (high:low or low:high; binomial probability = 0.0123).

**Conclusions:** The quadrant classification captures the essential variation in GERD subjects, aligns with phenotype groupings, reflects symptom burden (which phenotypes ignore), and maps more directly to possible treatment decisions. Given its simplicity and interpretability, a quadrant-based diagnosis and subsequent treatment choice may provide a pragmatic, evidence-based approach in routine clinical care.

## INTRODUCTION

The Lyon Consensus [1, 2] identified three distinct phenotypes of symptomatic gastroesophageal reflux disease (GERD) subjects: NERD, reflux hypersensitivity, functional heartburn, all defined in terms of percent time esophageal pH<4 and whether symptoms were associated with reflux events defined initially by a decrease in esophageal pH, and later by impedance [1, 2]. Having distinct diagnostic categories can facilitate diagnosis, treatment, and research; however, none of the Lyon phenotypes describe a relationship between esophageal acid exposure and symptom frequency – particularly in individual subjects [1, 2]. Moreover, the Lyon Consensus stated that the association of esophageal acid with symptoms is weak making the sole reliance on these measures problematic [1].

Because esophageal acid in GERD subjects is believed to be related to symptoms, the most common treatments involve reducing esophageal acid by inhibiting gastric acid secretion [3]. In the present analyses, we were interested in developing a method to relate symptom frequency to esophageal acid in individual GERD subjects in ways that might facilitate diagnosis, treatment, and research. Doing so has given rise to what we have defined as the “quadrant method”.

## METHODS

### Subjects

Subjects for the present analyses have provided data for previous analyses and the descriptions of subjects and some of the methods have been published previously (4).

All subjects had impedance–pH monitoring performed for 24 hours (Sandhill Scientific) after overnight fasting with gastric antisecretory agents having been discontinued for at least 7 days. The impedance–pH catheter was inserted with an esophageal pH sensor positioned 5 cm above the upper border of the lower esophageal sphincter, and a gastric pH sensor positioned 15 cm below the esophageal pH sensor. Six impedance channels were positioned 3, 5, 7, 9, 15, and 17 cm above the lower esophageal sphincter, respectively. The pH electrodes were calibrated following the instructions from the manufacturer and using their standard calibration solutions at pH 4 and pH 7. Because antimony electrodes, which are temperature sensitive, were used to record pH, the software provided by Sandhill to process pH recordings automatically adjusted all pH values for the difference between the calibration temperature 25°C and the recording temperature 37°C. Software provided by the manufacturer was also used to export pH data for every 4th second of the recording to a Microsoft Excel file. Values of pH below 0.5 were replaced with 0.5, and pH values above 7.5 were replaced with 7.5. Subjects were instructed to follow their usual daily routines and meals, but to avoid carbonated or acidic beverages. Subjects pressed an event marker to signal the meal periods, recumbent position, and symptoms of heartburn, regurgitation, or chest pain.

For the present analyses, subjects with a normal upper endoscopy were grouped using the Lyon consensus criteria for the three GERD phenotypes [1, 2]. Twenty subjects with normal esophageal acid exposure time (AET) (pH <4 for less than 4% of the 24-hour esophageal pH recording) plus a positive Symptom Index (SI) and a positive Symptom-Associated Probability (SAP) were assigned to the Reflux Hypersensitivity group. Twenty subjects with normal esophageal AET plus a negative SI and a negative SAP were assigned to the Functional Heartburn group. Twenty subjects with AET greater than 6% regardless of values for SI and SAP were assigned to the non-erosive reflux disease (NERD) group.

For this retrospective analysis of clinically indicated tests with no identifiable patient data, the Stanford University Institutional Review Board determined that this research does not involve human subjects as defined in 45 CFR 46.102(f) or 21 CFR 50.3 (g).

### Procedures

All individual pH recordings were processed as described in detail previously [4]. Interval esophageal acidity was measured for each pH recording from the beginning of the recording until the time of the 1st symptom, from the time of the 1st symptom until the time of the 2nd symptom and so on until the time of the last symptom. Interval acidity was calculated as the time-weighted average of the acid concentration expressed as mM.hr, and represents the area under the acid concentration-time curve. The median value for interval esophageal acidity was also calculated for each subject.

Most software used to process values for esophageal pH recording and symptom occurrence can export the data as text files, and it is a simple, straightforward process to then calculate values similar to those in the present study for an individual subject.

Median values for symptom frequency and esophageal acid were calculated for each GERD phenotype. In addition, for an individual phenotype, values for a particular measure from each subject were ranked from highest to lowest. For each measure, ranked values from one phenotype (e.g., phenotype A) were then compared to corresponding values from a second phenotype (e.g., phenotype B) to determine if the value from phenotype A was higher than the corresponding value from phenotype B. The sum of the differences is reported as A>B. The number of differences where values from phenotype B were greater than corresponding values from phenotype A is 20 minus the value for A>B. The sum of the differences was compared to chance difference by the binomial probability.

The major objective of our analyses is to examine the relationship between esophageal acid, and symptom frequency in individual GERD subjects.

#### Quadrant Figure

The Quadrant Figure examines relationships between pairs of measurements in the different phenotypes by plotting values for symptom frequency from an individual subject versus the corresponding value for median interval esophageal acid, from the same subject. Thus, each figure contains 20 pairs of values color-coded for each GERD phenotype or 60 pairs total.

The median value for a given measure was calculated for all 60 subjects combined, and two solid lines were added to the figure to indicate the median value for values on the X-axis and Y-axis. These solid lines divide each figure into four quadrants. We chose the median value because it is simple, robust to outliers and ensures equal-sized groups above and below each median line.

Values in the upper right quadrant in the figure are above the median value for both measures and are labeled “HIGH-HIGH”, and values in the lower left quadrant represents values that are below the median value for both measures and are labeled “LOW-LOW”. Values in the lower right quadrant are above the median value on the X-axis, but below the median value on the Y-axis and are labeled “HIGH-LOW”, while values in the upper left quadrant are below the median value on the X-axis, but above the median value on the Y-axis and are labeled “LOW-HIGH”.

#### Quadrant Table

The Quadrant Figure is accompanied by a table that lists the number of subjects from each GERD phenotype in each quadrant of the accompanying figure plus the value for the Chi-Square test for Independence that gives the probability that the observed distribution of the values in the table represents an independent association of the values. A low p-value from the test indicates that the distribution of phenotypes across the quadrants is not random and there is a significant association between phenotype and quadrant assignment.

#### Statistics

Statistical analyses were performed using GraphPad Prism 10.4.0. P-values were not adjusted for multiple comparisons, as the analyses were exploratory.

## RESULTS

Table 1 shows that symptom frequency in NERD subjects was not significantly different from that in reflux hypersensitivity subjects, but values in each phenotype were significantly higher than those in functional heartburn subjects. Table 1 also shows that esophageal acid in NERD subjects was significantly higher than that in reflux hypersensitivity subjects that, in turn, was higher than that in functional heartburn subjects. The Lyon Consensus reported that NERD subjects have esophageal acid exposure times above 6%, while both reflux hypersensitivity, and functional heartburn subjects have esophageal acid exposure times below 4% [1, 2]. Thus, the present analyses capture a difference in esophageal acid exposure that is not present in the Lyon Consensus classification.

**TABLE 1.**
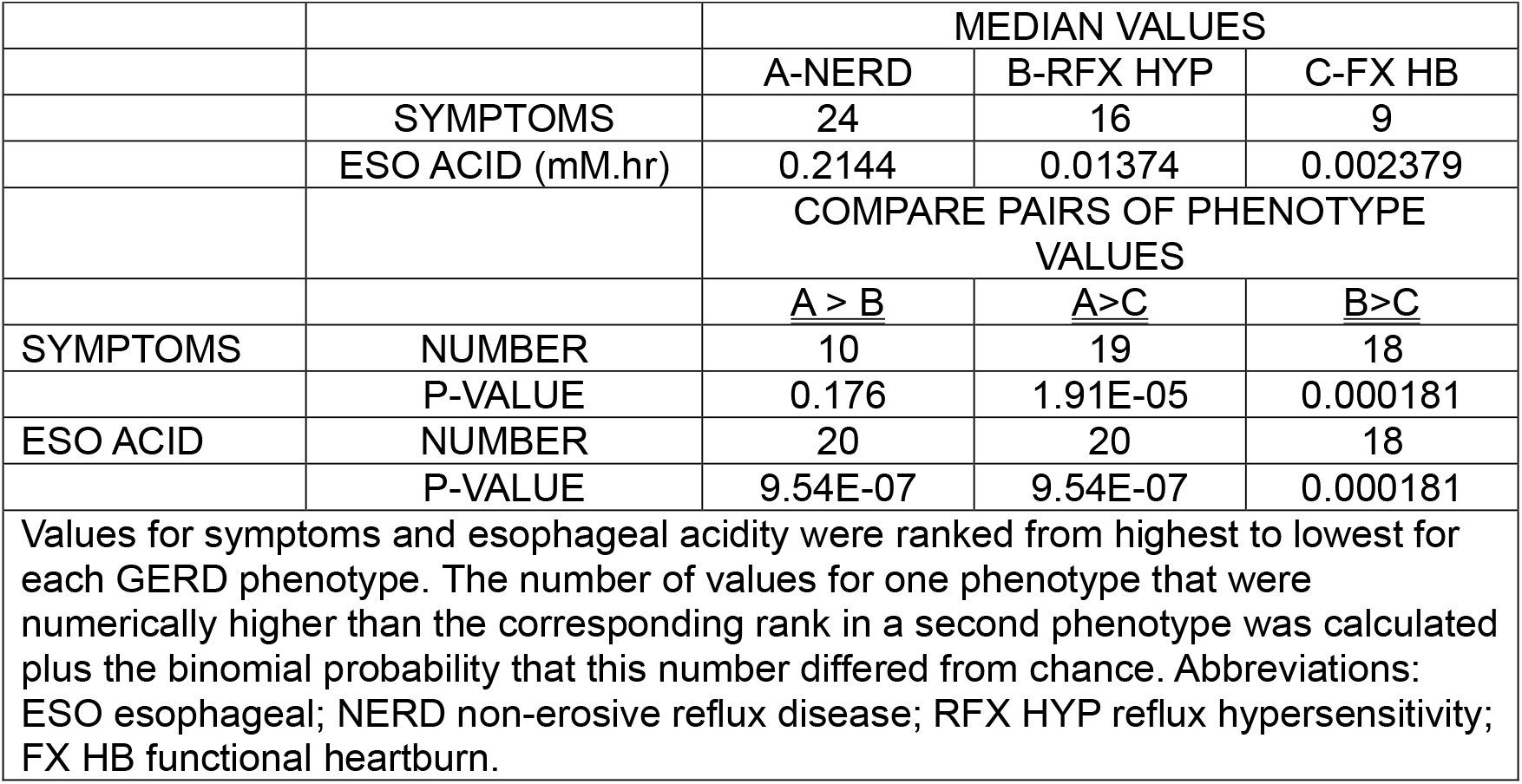
MEDIAN VALUES FOR SYMPTOMS AND ESOPHAGEAL ACID IN DIFFERENT GERD PHENOTYPES.

Figure 1 illustrates the marked variation in the relationship between symptom frequency and esophageal acid in 60 GERD subjects. Values from subjects from each GERD phenotype occur in each quadrant except for NERD subjects in the lower left quadrant (low: low) and functional heartburn subjects in the upper right quadrant (high: high). Figure 1 and Table 2 show that 85% of NERD subjects had high esophageal acid, as might be expected, but that 45% of subjects with reflux hypersensitivity and 20% of subjects with functional heartburn also had high esophageal acid.

**TABLE 2.** DISTRIBUTIONS OF VALUES FOR INTERVAL ESOPHAGEAL ACID AND SYMPTOMS IN DIFFERENT GERD PHENOTYPES.

|  |  |  |  |  |  |
| --- | --- | --- | --- | --- | --- |
| ESO ACID | SYMPTOMS | NERD | RFX HYP | FX HB | TOTAL |
| HIGH | HIGH | 9 | 2 | 0 | 11 |
| HIGH | LOW | 8 | 7 | 4 | 19 |
| LOW | HIGH | 3 | 9 | 7 | 19 |
| LOW | LOW | 0 | 2 | 9 | 11 |
|  | TOTAL | 20 | 20 | 20 | 60 |
| CHI-SQUARE test for Independence 17.20; DF 2; P=0.00018 |  |  |  |  |  |
| Data are from values in the quadrants illustrated in Figure 1. Abbreviations :ESO esophageal; NERD non-erosive reflux disease; RFX HYP reflux hypersensitivity; FX HB functional heartburn. The Chi-Square test was performed after combining the two values for high esophageal acid, and the two values for low esophageal acid in each column to avoid violating the assumptions of the Chi-Square test. |  |  |  |  |  |

**FIGURE 1.**
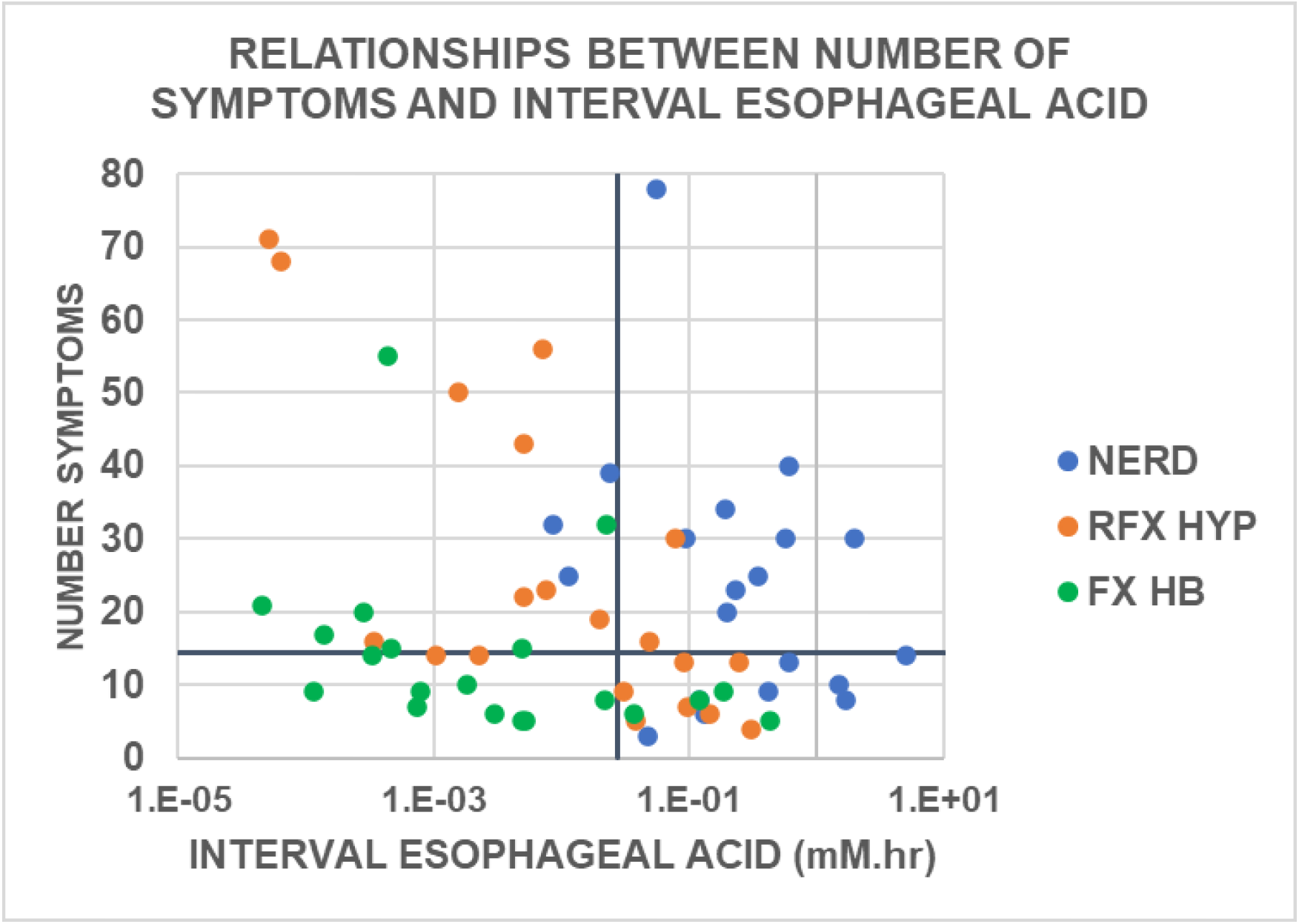
Relationships between interval esophageal acid and number of symptoms in different GERD phenotypes. The solid lines indicate the median of combined values from all three phenotypes (Symptoms 14.5; interval esophageal acid 0.0274 mM.hr.). Values above the horizontal solid line or to the right of the vertical solid line are labeled “high”, while values below the horizontal solid line or to the left of the vertical solid line are labeled “low”. Values are from 20 subjects in each phenotype. Abbreviations: RFX HYP reflux hypersensitivity; FX HB functional heartburn.

The Lyon Consensus does not report symptom frequency in the various phenotypes, but the quadrants in Figure 1 and Table 2 provide values for symptom frequency for each phenotype. The number of subjects with low symptom frequency in NERD, reflux hypersensitivity, and functional heartburn was 8, 9 and 13, respectively, indicating that at lease 40% of GERD subjects had low symptom frequency. In addition, functional heartburn subjects had the lowest values for symptom frequency, but the highest percentage of subjects with low symptoms.

Nearly all published literature in the GERD field specifies or implies a direct relationship between esophageal acid and symptoms. For example, the occurrence of GERD symptoms has been reported to be associated with increased esophageal acid exposure time during the time before the symptom [5, 6] and reductions in esophageal acidity with gastric antisecretory agents such as proton pump inhibitors (PPIs) reduce the occurrence of symptoms [1]. If there were a direct relationship between symptom frequency and esophageal acid in Figure 1, the points would fall on a diagonal going from the lower left to the upper right of the Figure.

The row totals in Table 2 illustrate the overall variation in relationships between symptom frequency and esophageal acid in GERD subjects in the present study. A majority of subjects (63%) had a discordant relationship between symptom frequency and esophageal acid that was significantly different from 50% (binomial probability = 0.0123). Of the 60 subjects, 19 had high esophageal acid and low symptom frequency, and 19 had low esophageal acid and high symptom frequency. A minority of subjects (37%) had a concordant relationship between symptom frequency and esophageal acid. Eleven subjects had high esophageal acid and high symptom frequency, and 11 had low esophageal acid and low symptom frequency.

- The high esophageal acid-high symptoms category reflects acid-driven symptoms in with 9 of the 11 subjects being NERD subjects.
- The high esophageal acid-low symptoms category indicates subjects that have a blunted response to esophageal acid or silent reflux that may reflect desensitization or something that inhibits the action of acid on the esophageal mucosa, and this category is dominated by NERD and reflux hypersensitivity subjects.
- The low esophageal acid-high symptoms category indicates subjects that have increased sensitivity to minimal esophageal acid exposure or another cause of symptoms besides esophageal acid, and this category is dominated by reflux hypersensitivity and functional heartburn subjects.
- The low esophageal acid-low symptoms category indicates subjects that have minimal or asymptomatic disease activity with 9 of 11 subjects being functional heartburn subjects. This category may represent non-GERD pathology.

The columns in Table 2 illustrate the variation in the relationship between esophageal acid and symptom frequency in the different GERD phenotypes.

- Seventeen NERD subjects had high esophageal acid with 9 having high symptom frequency acid and 8 having low symptom frequency.
- Sixteen functional heartburn subjects had low esophageal acid with 7 having high symptom frequency and 9 having low symptom frequency.
- Of sixteen reflux hypersensitivity subjects, 7 had high esophageal acid and low symptom frequency, while 9 had low esophageal and high symptom frequency.

These results illustrate that each phenotype preferentially exhibits a distinct combination of esophageal acid exposure and symptom frequency patterns, suggesting differing underlying pathophysiologic mechanisms. The P-value of 0.00018 from the Chi-Square test given in Table 2 indicates that there is a significant correlation between quadrant distributions and GERD phenotype [7]. As a result, the quadrant patterns reflect core diagnostic features and reinforce the validity of quadrant-based classification as clinically meaningful

## DISCUSSION

Our data illustrate that the quadrant classification captures the variation in the relationship between interval esophageal acid and symptom frequency among GERD subjects. The low P-value from the Chi-Square test for Independence indicates that the variation in the relationship between interval esophageal acid and symptom frequency identified by the quadrant method also correlates highly with that among the different GERD phenotypes identified in the Lyon Consensus subjects [7]. Thus, the quadrant method captures the essential variation in GERD subjects. It aligns with phenotype groupings, reflects symptom burden (which phenotypes ignore), and maps more directly to possible treatment decisions. Overall, the quadrant-based analysis is useful for clinical inference, while the phenotype-based analysis aids in mechanistic understanding. Given its simplicity and interpretability, quadrant-based diagnosis can provide a pragmatic, evidence-based complement to traditional phenotyping in routine clinical care.

The findings in the Lyon Consensus analyses that subjects with Reflux Hypersensitivity or Functional Heartburn have GERD symptoms but normal esophageal acid exposure time have led to the inference that symptoms may be caused by esophageal acid hypersensitivity [8-11]. Our data, however, show that only a minority of subjects with these phenotypes have low esophageal acid coupled with high symptom frequency - 45% of subjects with reflux hypersensitivity and 35% of subjects. These results indicate the potential value of quadrant analyses to identify GERD subjects for possible esophageal acid hypersensitivity or non-acid causes of symptoms.

A major finding in our study is that most symptomatic GERD subjects (63%) show a discordant relationship between symptom frequency and interval esophageal acidity, i.e., either high esophageal acid and low symptom frequency or low esophageal acid and high symptom frequency. These results were surprising because of the general belief that in most subjects with symptomatic GERD, there is a direct relationship between esophageal acid exposure and symptom frequency, and treatment should be primarily with agents that reduce esophageal acid exposure by inhibiting gastric acid secretion. The quadrant method illustrates how GERD subjects with such a discordant relationship between symptom frequency and esophageal acid exposure can be readily identified and treated differently.

Approximately forty percent of GERD subjects have been reported to fail treatment with a PPI, and multiple explanations have ben proposed for these failures [12]. A common recommendation has been to conduct impedance-pH measurements to determine the presence of reflux and its association with symptoms [12]. These measurements have typically measured time esophageal pH<4 plus the possible association of symptoms with reflux episodes. They have not, however, included measurements of the relationship between esophageal acid and symptom frequency. The quadrant method allows evaluation based on actual symptom burden and accompanying esophageal acid exposure, not just their temporal association. As a result, it maps more directly to possible diagnosis and treatment.

## Data Availability

All data produced in the present study are available upon reasonable request to the authors

## ACKNOWLEDGMENT

The authors thank Dr. Daniel Sifrim, Director of Upper GI Physiology Unit, Royal London Hospital, for providing the impedance–pH records used for the present analyses.

## REFERENCES

1. Gyawali CP, Kahrilas PJ, Savarino E, et al. Modern diagnosis of GERD: the Lyon Consensus. Gut. 2018; 67:1351–1362. doi: 10.1136/gutjnl-2017-314722.

2. Ghisa M, Barberio B, Savarino V, et al. The Lyon Consensus: Does It Differ From the Previous Ones? J Neurogastroenterol Motil. 2020;26:311–321. doi: 10.5056/jnm20030.

3. Yadlapati R, Gyawali CP, Pandolfino JE on behalf of the CGIT GERD Consensus Conference Participants. AGA Clinical Practice Update on the Personalized Approach to the Evaluation and Management of GERD: Expert Review. Clinical Gastroenterology and Hepatology 2022;20:984–994.doi:10.1016/j.cgh.2022.01.025.

4. Gardner JD. The relationship between esophageal acidity and symptom frequency in symptomatic nonerosive gastroesophageal reflux disease. Physiological Reports, 10, e15442. doi: 10.14814/phy2.15442.

5. Beedassy A, Katz PO, Gruber A. Prior sensitization of esophageal mucosa by acid reflux predisposes to reflux-induced chest pain. J Clin Gastroentrol 2000; 31:121–124. doi: 10.1097/00004836-200009000-00006

6. Bredenoord AJ, Weusten BLAM, Curvers WL, et al. Determinants of perception of heartburn and regurgitation. Gut 2006; 55:313–318. doi: 10.1136/gut.2005.074690

7. Sheskin DJ. Handbook of parametric and nonparametric statistical procedures. Boca Raton: Chapman & Hall/CRC, 2007: 619–740.

8. Bredenoord AJ. Mechanisms of reflux perception in gastroesophageal reflux disease: a review. Am J Gastroenterol 2012; 107:8–15. doi: 10.1038/ajg.2011.286

9. Howard PJ, Maher L, Pryde A, et al. Symptomatic gastroesophageal reflux, abnormal oesophageal acid exposure, and mucosal acid sensitivity are three separate, though related aspects of gastro-oesophageal reflux disease. Gut 1991; 32:128–132. doi: 10.1136/gut.32.2.128

10. Trimble KC, Pryde A, Heading RC, et al. Lowered oesophageal sensory thresholds in patients with symptomatic but not excess gastro-oesophageal reflux. Evidence for a spectrum of visceral sensitivity in GORD. Gut 1995; 37:7–12. doi: 10.1136/gut.37.1.7

11. Rodriguez-Stanley S, Robinson M, Earnest DL, et al. Esophageal hypersensitivity may be a major cause of heartburn‥ Am J Gastroenterol 1999; 94:628–631. doi: 10.1111/j.1572-0241.1999.00925.x

12. Rettura F, Bronzini F, Campigotto M, et al. Refractory gastroesophageal reflux disease: A management update. Front Med 8:765061. doi: 10.3389/fmed.2021.765061.

